# Body surface electrical recordings detect alterations in colonic motility and heart rate variability in irritable bowel syndrome patients

**DOI:** 10.64898/2026.06.02.26354686

**Authors:** Jonathan C. Erickson, London Paige, Jasmine Gipson, Natalie Gresham, Phil G. Dinning

**Affiliations:** Department of Physics and Engineering, Washington and Lee University, Lexington, VA, USA; Department of Surgery, University of Auckland, New Zealand; Department of Surgery and Gastroenterology. Flinders Medical Centre. College of Medicine and Public Health, Flinders University, Adelaide, Australia

**Author notes:** Correspondence: JCE.

**Keywords:** colonic motility, body surface colonic mapping (BSCM), disorders of gut-brain axis (DGBI), heart rate variability (HRV), Irritable bowel syndrome (IBS)

## Abstract

Irritable Bowel Syndrome (IBS) is a highly prevalent, commonly diagnosed gastrointestinal disorder of gut-brain interaction (DGBI) that causes substantial physical, psychological, and financial burden. The role of abnormal motility and altered autonomic nervous system function, and their interplay, remains to be fully understood. Here we present a non-invasive method using body surface electrical recordings to concurrently quantify meal-response colonic motility and heart rate variability (HRV). We demonstrate the practical utility of this new technique in a pilot study comparing colonic motility and autonomic nervous system (ANS) function in IBS patients (n=14) and healthy controls (HC; n = 22). The study protocol included a 2-3 hr body-surface electrical recording with 60-90 minutes each of pre- and post- meal epochs. Colonic motility was markedly increased in the subset of IBS patients experiencing moderate-to-severe symptoms during the study, compared to IBS no or mild symptom groups and healthy controls. HRV metrics in IBS patients showed substantial baseline shifts with decreased vagal and increased sympathetic input, with blunted autonomic meal responses compared to HC. Newly introduced dynamic trajectory maps revealed pronounced colon motility-vagal dysregulation in high symptom IBS patients but not mild or no symptom groups. These results indicate altered autonomic-motility interaction as a potential mechanism of symptom genesis in IBS patients. This technology platform offers an easy-to-apply, non-invasive tool for larger scale investigations of gut and autonomic nervous system function in healthy and gastrointestinal disease cohorts.

**NEW & NOTEWORTHY:** This work presents a novel method for investigating interactions between colonic motility and autonomic function using non-invasive body-surface electrical recordings. We demonstrate practical utility by comparing meal-induced responses in an irritable bowel syndrome (IBS) patient cohort and healthy controls. Significant parasympathetic-motility dysregulation was observed in IBS patients experiencing a high symptom burden. Our new method overcomes limitations of invasive measurement techniques, offering new possibilities to investigate mechanisms of gut-brain interaction in health and disease.\

## INTRODUCTION

Irritable bowel syndrome (IBS) impacts an estimated ∼11% of the world population (1, 2) with a substantial impact on quality of life and high social and economic burden (3–5). As a disorder of gut-brain interaction (DGBI), IBS is characterized by chronic abdominal pain related to defecation and changes in stool consistency. Its heterogeneous pathophysiology is hypothesised to be primarily driven by altered gut motility and autonomic nervous system (ANS) dysfunction (6–8). The ANS serves as the critical bidirectional communication link between the brain and the gut, modulating the enteric nervous system through sympathetic and parasympathetic pathways (9, 10). Consequently, ANS regulation in IBS patients is a significant area of clinical interest (11–14).

ANS function and regulation are typically assessed via Heart Rate Variability (HRV), a non-invasive proxy measure. This metric has been utilized to investigate physiological differences between patients with IBS and healthy controls, as well as variations across IBS subgroups and levels of symptom severity (Ali & Chen, 2023). Studies have shown that patients with severe IBS symptoms often exhibit altered HRV profiles, including vagal withdrawal or sympathetic dominance during orthostatic responses (15). HRV metrics were consistently altered in women with severe-IBS symptoms, but not those patients with a lower symptom burden (16). Notably, in IBS patients, symptomatic periods preceding defecation were associated with decreased parasympathetic and increased in sympathetic activity (17).

Altered colonic motility is also shown in IBS. Mucosal electrical recordings, showed that IBS patients had a marked increase in ∼3 cpm (cycles per min) contractions following cholecystokinin and pentagrastrin stimulation (18). An increase in the propulsive high-amplitude propagated contractions (HAPCs) and diminished non-propagating contractions in the distal colon, was shown in patients with functional diarrhea (19). High-resolution manometry in IBS-D also captured a significant reduction in sigmoid colon cyclic motor pattern (20) HAPCs have also been associated with abdominal pain in IBS (21). Some evidence suggests a correlation between postprandial motility and vagal withdrawal in IBS (22) while studies of refractory chronic constipation have identified a combination of mild vagal impairment, high sympathetic tone, and reduced HAPC amplitudes (23). In healthy individuals, HAPCs appear to synchronize with a shift toward parasympathetic dominance, suggesting that the loss of this autonomic coordination may drive IBS symptoms. (24).

A major barrier to advancing this field is that traditional manometry is highly invasive, resource-intensive, and requires hospitalization, making it impractical for large-scale clinical use. Overcoming these methodological constraints is essential for better characterizing the diverse pathophysiology of IBS. This study proposes Body Surface Colonic Mapping (BSCM) as a feasible, non-invasive, and accessible alternative for the simultaneous assessment of colonic motility and HRV parameters, potentially offering a scalable solution for integrated gut-brain research.

## METHODS

### Ethics Statement

Data was collected with informed consent of the participants and with approval from the Institutional Review Board for Research with Human Subjects at Washington and Lee University (IRB.201718.034).

### Inclusion and exclusion criteria

All study participants had to be at least 18 years of age with a BMI < 30. Volunteers accepted into the healthy controls cohort had a normal bowel habit, with no symptoms of IBS and no adverse gastrointestinal symptoms in the 48 hrs leading up to the test. IBS patients (n=14) accepted into the study met Rome IV criteria (25). Exclusion criteria applied to candidates with known dermal contact allergies; cases of confirmed or suspected pregnancy; and major comorbidities such as prior gastrointestinal surgery or spinal injury.

Participants with diarrheal predominant (IBS-D; 11/14 = 78.6%) and constipation predominant (IBS-C; 3/14 = 21.4%) subtypes were pooled for analyses. The rationale supporting this choice was: 1) prior work indicated no significant difference in HRV parameters between IBS-D and IBS-C subtypes (26, 27), although contrary findings have also been reported (28); 2) mechanisms of altered colonic motility in IBS-D and IBS-C may overlap—i.e., attenuated retrograde motility and amplified antegrade activity may lead to diarrhea, and vice versa for constipation; and 3) maximize the size of the IBS cohort to achieve the broader aim of this work establishing feasibility of a new method for analyzing colonic motility and ANS profiles relative to healthy controls.

### Study protocol and data acquisition

Prior to the study, participants were asked to fast overnight, refrain from caffeinated and alcoholic beverages, and moderate to heavy exercises. These conditions were used to minimize colonic activity prior to the start of the study (29). Our study protocol included a 60–90-minute recording period, before and after a meal (2-3 hours total). To avoid undue stress on IBS patients, participants were allowed to eat a meal they considered to be of standardized size and average caloric content for their daily diet (∼750kcal).

Prior to the start of the recording, study participants applied Nuprep® abrasive skin prepping gel to their abdomen. We affixed Ag/AgCl, 22 × 44 mm adhesive cutaneous electrodes in either a 5 × 6 or 4 × 8 (row × column) configuration, targeting the rectosigmoid region of the colon (30, 31). The area of coverage was dependent on the participants’ size and shape but generally spanned the abdominal surface horizontally between the left iliac crest spanning toward or across their midline, and vertically from the umbilicus down to the upper region of the symphysis pubis (Figure 1). The reference electrodes were placed on the right iliac crest. A modified cardiac stress test elastic net was used to secure electrode contact and reduce motion artifacts.

**Figure 1.**
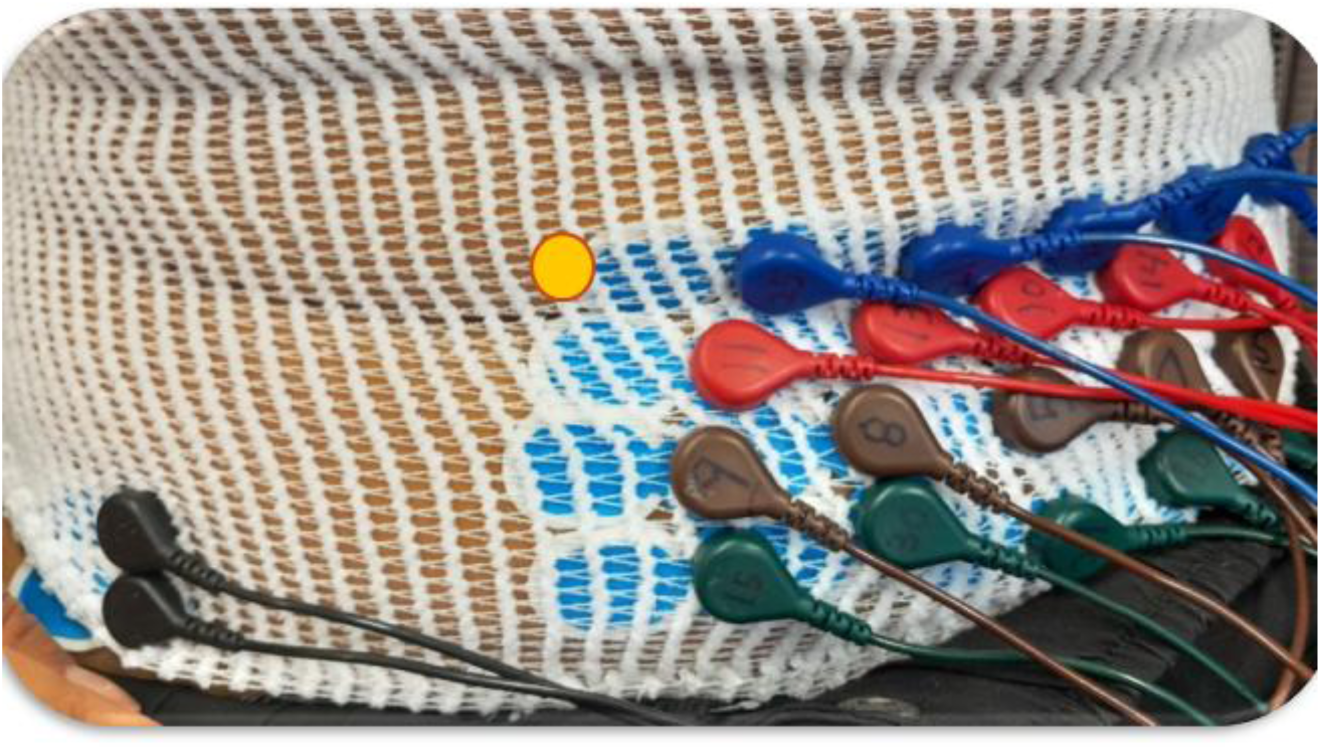
Example body surface electrode array (20 channels, 4x5 configuration) positioned over lower left abdomen. Ground and reference electrodes (black leads) were positioned near the right hip iliac crest. A modified cardiac stress test elastic net was used to help secure electrodes and leads in place. Orange circle marks the position of the umbilicus.

The Intsy multichannel bioamplifier module (32, 33) recorded body surface electrical signals at a sampling rate of 100 Hz, enabling analysis of colonic motility and cardiac activity to determine HRV parameters. The Intsy system was previously validated for recording electrical signals of colonic origin with frequency components as low as approximately ∼1.2 cpm (lower cutoff of 0.02 Hz).

During the recordings, subjects rested in a comfortable reclined position (torso angle ∼15-30 deg) during the pre- and post-prandial epochs. Some subjects were temporarily inclined to ∼45 deg position to facilitate comfort during meal intake, returning to the originally reclined position when finished eating.

### Symptom Burden: Qualitative Reports and Categories

IBS subjects were asked to keep a written log of notable GI events that occurred throughout the study, such as sensation of pain or bloating, gassiness, flatus and urge to defecate or defecation. They were also asked to provide an oral and written assessment of their symptoms experienced during the study. Based on these responses, the total symptom burden was subjectively assigned to one of three categories: ‘none’ (n=6), ‘mild’ (n=5), ‘moderate to severe’ (n=3). Example remarks provided by subjects in each of the 3 categories were as follows:

> None: *‘feeling comfortable, essentially no discomfort, gassiness or other symptoms throughout the study today.’*
>
> Mild: *‘General feeling of bloating, increased sensation/especially prominent after meal. Also experienced light wave of nausea.’*
>
> moderate to severe: *‘bloating and significant cramping throughout post-meal period, progressively worse and severe at times; multiple passing waves of strong urge but withheld toileting.’*

### BSCM Signal Processing: colonic motility index

Colonic motility was extracted from the electrode data following a signal processing pipeline that has been previously detailed (30, 31, 34) Briefly, data were processed in 4 main steps:

1. Remove baseline wander with a moving median filter (temporal window of 20 sec window)
2. Apply a 4-10 cpm digital band pass filter (zero-phase, Butterworth, 2^nd^ order) to isolate colonic components and attenuate noise and interference sources (including respiration and gastric activity at ∼ 3cpm). This frequency range was found to be optimal for detecting colonic meal responses in previous work (34).
3. Remove large amplitude transient waveforms attributed to motion using a temporal Wiener filter with adaptive noise calculation parameters (30 second local variance and 300 sec global variance time windows)
4. Compute the continuous wavelet transform and extract the colonic motility index (MI) from the spectrogram.

MI changes stimulated by meal-intake were analyzed using 60 min duration pre- and post-prandial epochs (120 minutes total), with the meal start synchronized at t = 0. Pre-and post-meal differences in MI were quantified using two metrics. The meal response ratio (MRR) measures the maximum change in postprandial MI relative to the pre-meal baseline. The amplitude percent difference (APD) measures the difference in average post-meal epoch MI relative to pre-meal baseline.

While the primary results reported here are for the 4-10 cpm band (step 2), for case study analyses colonic motility was also assessed using the 0.5-10 cpm range, motivated by a recent report demonstrating detection of HAPCs using this frequency band (35).

### R-Peak Detection and HRV metrics

Automated R-peak detection is a prerequisite for computing HRV metrics. All data was processed in custom MATLAB software. We selected a subset of 3 electrode channels for analysis meeting the criterion of having high signal quality via visual inspection—i.e. clearly identifiable cardiac waveforms for the duration of the study. We employed a modified Pan-Tompkins algorithm to detect and timestamp R-peaks. Pre-processing steps included removal of baseline wander (moving median filter, 20 s window) and digital filtering with a 5-25 Hz pass band (Butterworth, 2^nd^ order, zero phase) to isolate the QRS complex waveform and suppress P and T waveforms (36). In place of the traditional derivative squared signal, we computed the Teager-Kaiser Energy Transform because it amplifies large fast transients of the R-peak, making them easier to detect in the subsequent decision making steps (37).

The initial R-peak timestamps were used to calculate the sequence of interbeat intervals (IBI). The IBI intervals were then cleaned in a two-step process that 1) removed outliers deviating more than 3x the MAD (median of the absolute deviation) from the median value (Matlab function *isoutlier* using the Grubbs method); and 2) heuristic thresholding to eliminate IBI that falling outside of the range 0.5 – 2.5 s. This corresponds to a heart rate of 24-120 bpm, physiologically plausible values for subjects in a resting experimental condition. The instantaneous heart rate corresponding to the *k*_th_ interval was computed as *HR[n] = 1/(IBI[k] – IBI[k-1]*).

We computed a total of three HRV parameters as proxy measures of autonomic activity, summarized in Table 1. RMSSD serves as a measure of parasympathetic input and is associated with high frequency power band (Schaffer, F., Ginsber, J. P., 2017). SI serves as a measure of sympathetic input, is associated with the low-frequency power band, and was computed according to (38). The two main branches of the ANS can work in concert or in opposition, thus the autonomic ‘balance’ was quantified as the ratio of SI/RMSSD following (24). Prior work has indicated these time-domain HRV parameters are optimal for analyzing HRV to assess autonomic responses relation to colonic motility (24), especially in IBS patients (11).

**Table 1.**
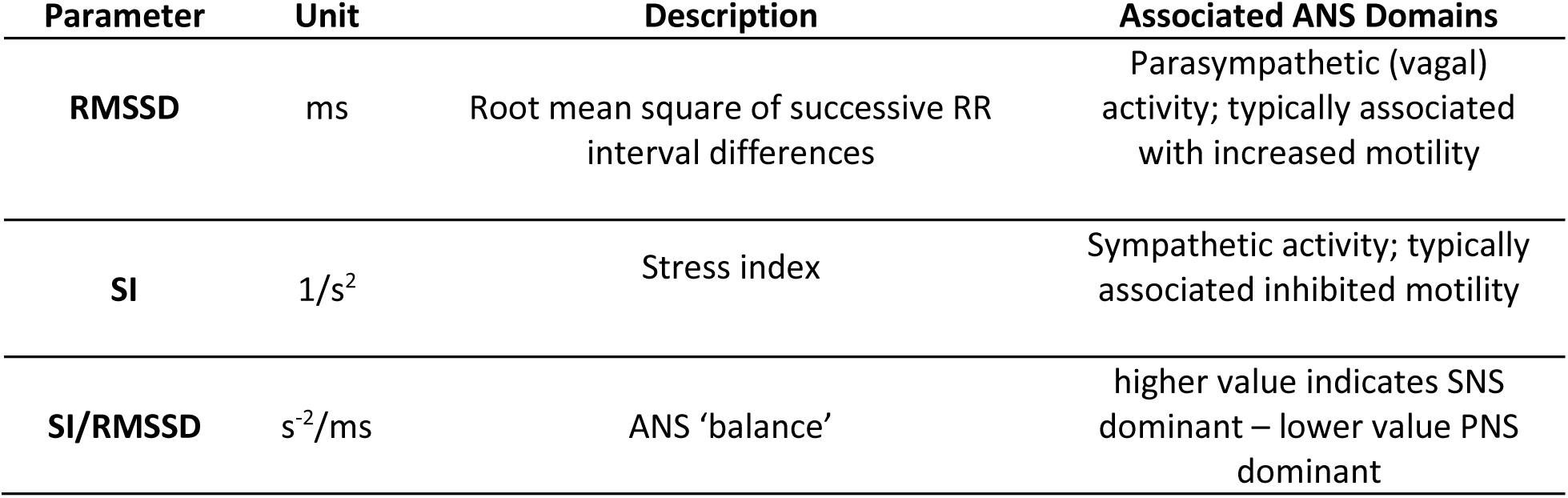
HRV parameter summary.

All HRV parameters were computed over the duration of the study using blocks of 100 successive IBIs, equivalently ∼2 min data windows for an average heart rate of ∼70 bpm. The values reported in this paper are the median HRV metric values across the 3 selected channels.

### Colonic motility-autonomic input associations: dynamic profile analysis

The association (or joint reactivity) between colonic motility and autonomic inputs may change over time, namely in response to the meal stimulus in the present study. To visualize and quantify these dynamic associations, we plotted 2-D trajectories of colonic motility index versus a selected HRV parameter, using the cohort-wide median for both measures. A stable relation would appear as a high-density of points confined to—in ‘tight-orbit’--around a small region in the 2-D plane. By contrast, dysregulated associations would manifest as more diffusely distributed points with relatively chaotic trajectories covering a wider area in the 2-D plane. Distribution and area coverage was further assessed by computing the pointwise distance to the trajectory centroids for the pre and postprandial periods.

The major axis of the trajectory relative to the vertical was defined by the line connecting the centroids. Bivariate kernel smoothed probability density maps were also generated from the set of points defining the colonic motility-HRV trajectory.

### IBS cohort analysis stratified by symptom burden

To discern potential contributions of abnormal motility, dysregulated autonomic response, and interactions thereof in driving IBS-related symptoms, our general analysis approach was to compare results for healthy controls to the subsets of IBS subjects stratified by symptom burden (none, mild, moderate/severe). The cohort sizes were healthy controls = 22; IBS no symptom = 6; IBS mild symptom = 5; IBS moderate/severe symptom = 3.

For motility-autonomic dynamic trajectory analyses, we accounted for the much larger size of the healthy control cohort by computing 30 subsamples of n = 6 randomly selected subjects. The core idea was to avoid biases in averaging effect inherent with larger cohort sizes. The distribution of correlation coefficients for the bivariate kernel smoothed density maps compared these subsets to IBS symptom subgroups as well as the full cohort of healthy controls. (See Supplemental Material for full details.)

### Statistical Analysis

Statistical analyses were performed using MATLAB (R2024a, MathWorks, Inc.). Given the relatively small and unequal sample sizes (*n_controls_*=22, *n_IBS_*=14), data were screened for normality using the Shapiro-Wilk test. Due to the presence of significant skewness and outliers in several HRV metrics (particularly SI/RMSSD and SI), both parametric and non-parametric descriptive statistics are reported (Median ± MAD and Mean ± SD) to provide a comprehensive view of the distribution.

Effect sizes comparing distribution means between IBS groups to healthy controls was calculated using Hedges’ g with correction term applied to account for the bias inherent in small sample sizes. Confidence intervals (CIs; 5-95%) for Hedges’ g were generated using bootstrap resampling (n_boot_ = 10000). An effect was considered statistically significant at the 0.05 level if the 5-95% CI did not cross the zero (null) line.

## RESULTS

### Cohort Demographics

Cohort demographics for HC and IBS cohorts are summarized in Table 2. BMI was well matched, while IBS patients were slightly older and the gender composition was reasonably similar.

**Table 2:**
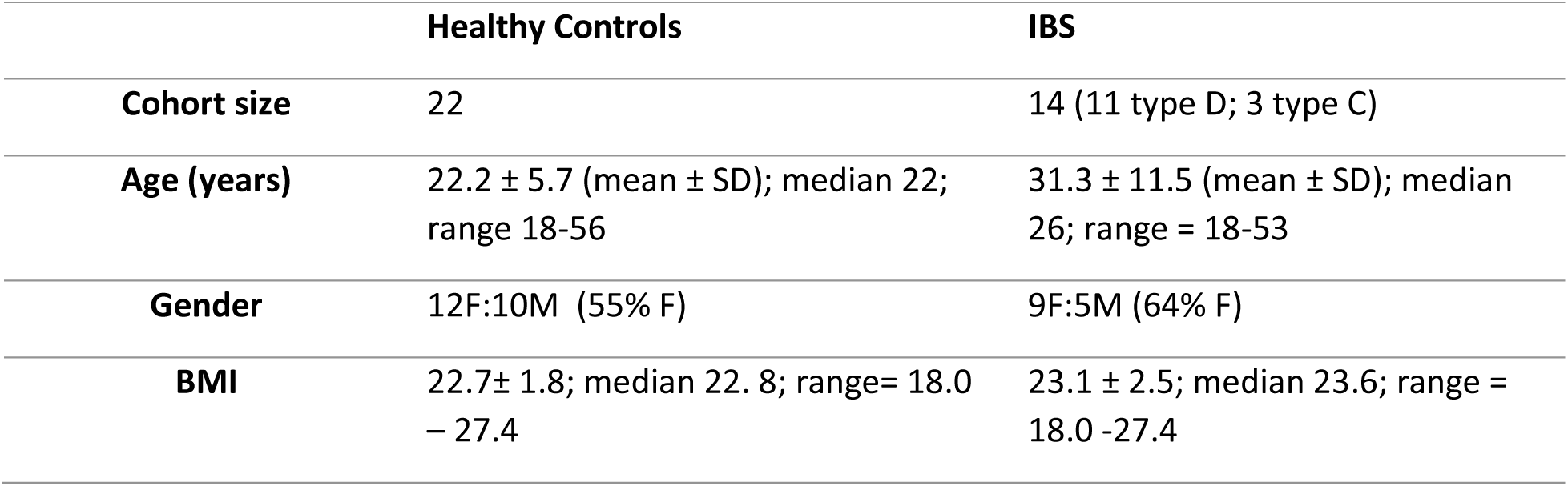
Cohort demographic summary.

### BSCM motility: IBS symptom burden subsets vs healthy controls

The averaged colonic motility for the baseline and meal response is shown in Figure 2. The IBS patients reporting no symptoms exhibit a striking similarity to healthy controls (Lin’s concordance correlation coefficient *ρ_c_* = 0.76, Pearson correlation = 0.84; Figure 2b). IBS patients with mild symptoms show an early and late increase in motility (*ρ_c_* = 0.53; Figure 2c). The MI for IBS patients experiencing moderate-to-severe symptoms exhibited large amplitude deviations compared to the other groups (*ρ_c_* = 0.55; Figure 2d).

**Figure 2.**
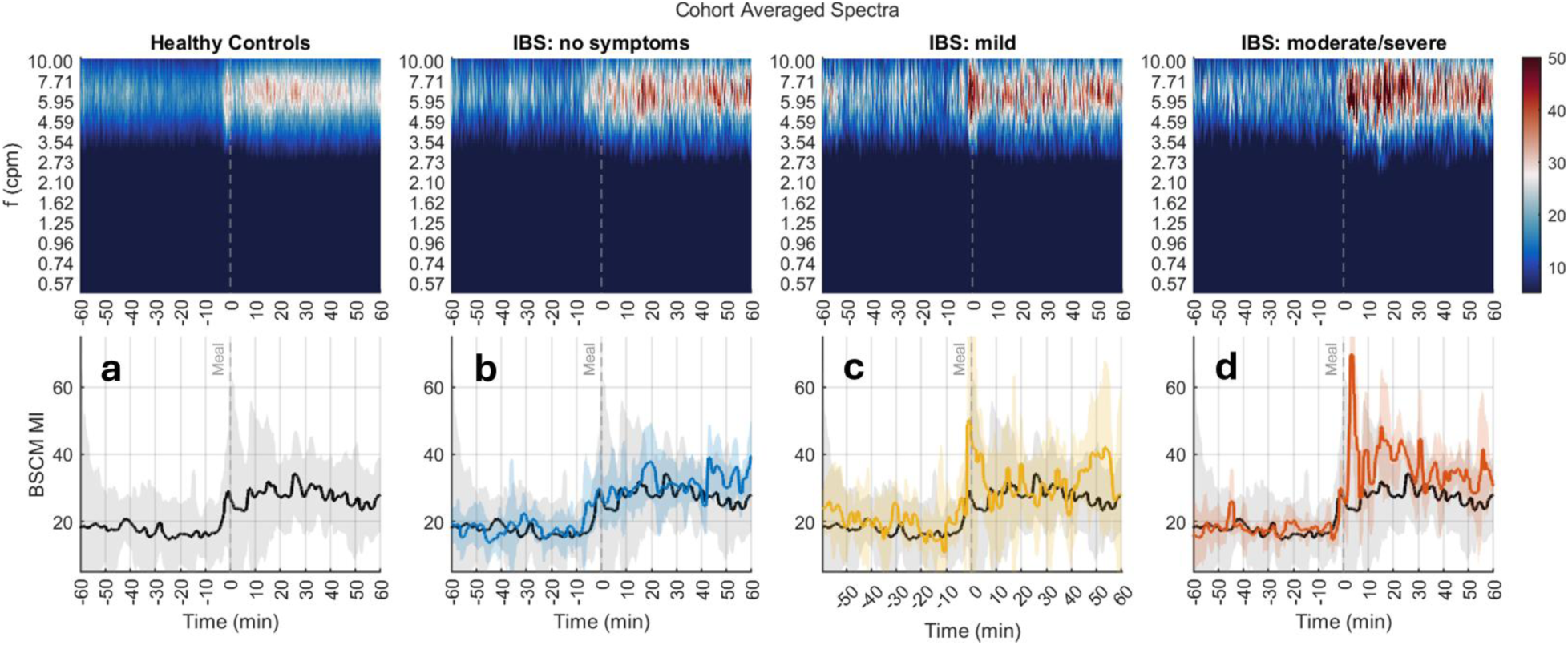
Cohort Averaged BSCM motility spectrograms and motility index (MI) vs time comparing healthy controls to IBS symptom burden subsets. The traces in the bottom row indicate the cohort median value and the back shading indicates the variance. The MI for healthy controls (black line; gray back shading) is duplicated in the IBS subgroup panels for ease of comparison.

The corresponding meal response motility metrics across the 4 groups are summarized in Figure 3 with summary statistics provided in Supplemental Material Table S1. In this pilot study, meal-induced colonic motility in the IBS no symptoms group was similar to healthy controls and then increased with IBS symptom severity. Dramatic differences noted in the moderate/severe symptom group.

**Figure 3.**
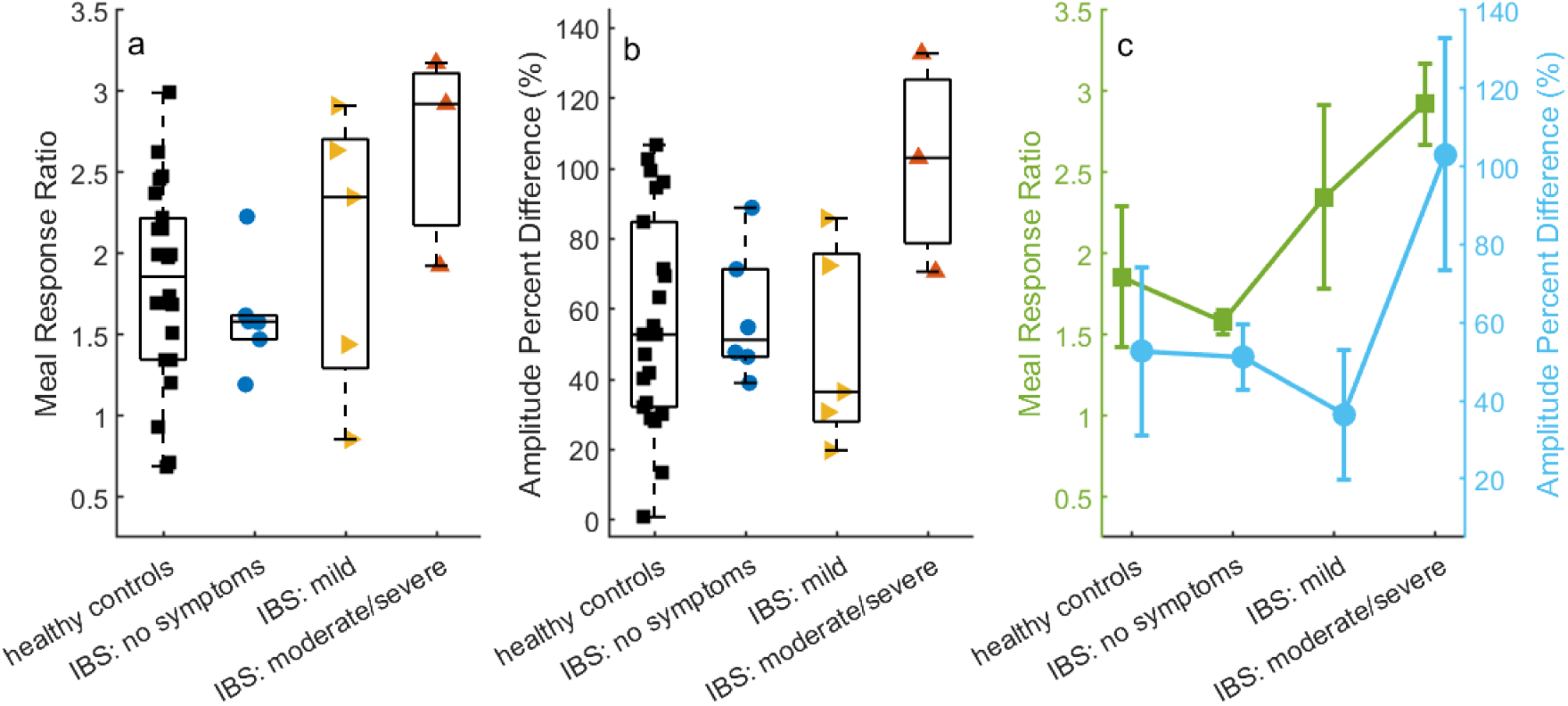
Comparison of meal response MI metrics for each group. **a**: meal response ratio indicates the maximum change in MI induced by the meal relative to baseline. Meal response ratio increases with IBS symptom severity; **b**: change in time-averaged amplitude post- vs pre-meal. The IBS moderate/severe symptoms cohort shows marked increases relative to the other 3 groups; **c**: median +MAD summary for meal response ratio (green) and amplitude percent difference (blue).

The meal response ratio (MRR) increases linearly with IBS symptom severity (Figure 3a and 3c). The IBS no symptoms group and healthy controls exhibited similar values (1.58 ± 0.07 vs 1.86 ± 0.44; median ± MAD). The MRR for the IBS moderate/high symptom group (2.92 ± 0.25) was 85.0% higher compared to healthy controls and their distributions were significantly different (Wilcoxon rank sum test p-value = 0.048). The IBS mild symptom group response ratio values (2.35 ± 0.56) were higher than patients without symptoms and healthy controls but did not differ significantly from either (Wilcoxon p = 0.54 and 0.47, respectively).

The Amplitude Percent Difference (APD) followed similar trends (Figure 3b and 3c). The IBS moderate/high symptoms group exhibited ∼2x larger maximal MI increases (103 ± 29.7%) compared to the other 3 groups (52.7 ± 21.6%; 51.2 ± 8.5%, 36.4 ± 16.7%, respectively for healthy controls, IBS no and mild symptoms). The APD values for IBS moderate/severe symptom group were significantly different compared to healthy controls (Wilcoxon p value = 0.04). There were no other significant differences shown.

### HRV metric comparisons

Group median-averaged HRV metrics and HR comparing each IBS symptom group to healthy controls are illustrated in Figure 4 and summarized in Supplementary Information Table S2. Compared to healthy controls, IBS no and mild symptom groups exhibited a lower premeal baseline RMSSD (63.9 ms for HC vs 38.2 and 43.4 ms for IBS no and mild symptoms, respectively) and a higher SI (51.6 for HC vs 110.2 and 114.8 s^-2^). RMSSD responsiveness to the meal was attenuated for both IBS groups relative to controls (ΔRMSSD = -3.0 and -10.3 ms for IBS groups vs -12.9 ms for controls) .

**Figure 4.**
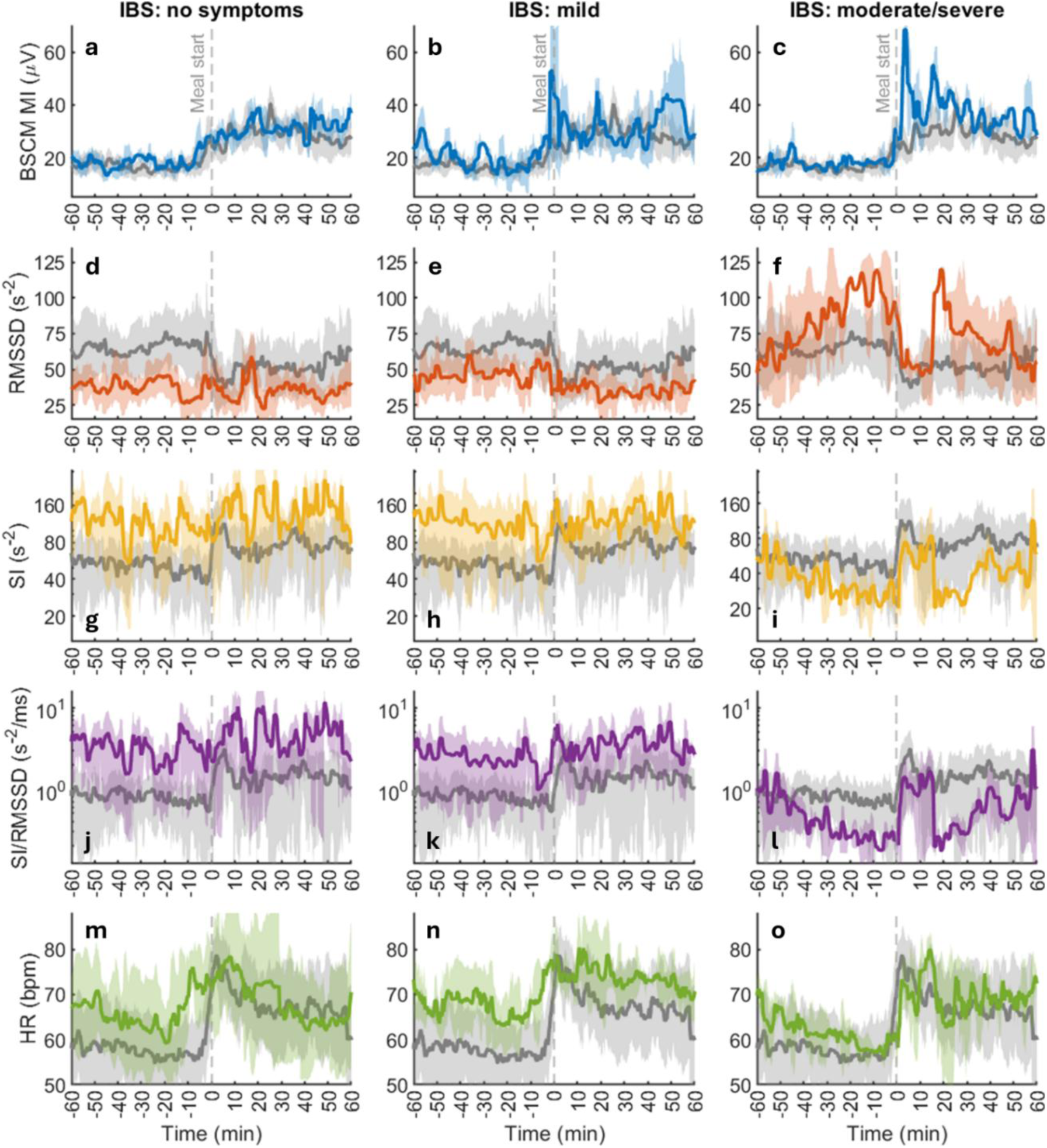
Cohort averaged BSCM motility and HRV metrics in IBS patient groups vs healthy controls. Columns are organized according to IBS symptom severity; separate metrics are organized in rows. Results for controls are shown as dark gray in all cases. Note log-scaled axis for SI and SI/RMSSD to facilitate visualization across orders of magnitude. RMSSD: IBS no and mild/moderate symptoms indicate decreased vagal tone with attenuated meal response. IBS moderate/severe symptoms showed markedly different RMSSD dynamics. SI: IBS patients for low and mild/moderate show increased sympathetic tone, with decreased responsivity to meal stimulus. Moderate/severe IBS group shows markedly different SI meal response dynamics. SI/RMSSD indicates ANS balance is biased toward sympathetic drive in the first two IBS groups compared to controls, while a bias toward vagal drive was observed in the moderate/high symptom group.

The IBS moderate/severe symptoms group demonstrated a dramatic increase in RMSSD over healthy controls during baseline, followed by a sharp, pulsatile change during meal ingestion followed by post-meal decrease returning to the initial pre-meal value by the end of the analysis epoch (Figure 4f). The SI exhibited similar dynamics, but in the opposite direction, ramping down pre-meal and ramping back up during the post-meal period (Figure 4i).

The SI/RMSSD ratio further highlighted the difference in HRV dynamics, which indicates a sympathetic shift in autonomic balance for the no and mild IBS symptoms groups. The moderate/severe symptoms IBS group exhibited a dysregulated vagal shift (Figure 4l).

The baseline HR for the no and mild symptom IBS groups was ∼10 bpm higher compared to controls, with a more modest and time-varying difference observed in the moderate/severe symptoms group (Figure 4m-o). Post-meal peak heart rates were similar (∼75 bpm) across all groups, however the post-meal HR remained high in the IBS mild symptoms group, while all others showed decreases. For the IBS no and mild symptoms group, the HR increase begins about 5-10 min earlier (prior to the meal), which could indicate anticipatory stress.

Subject-specific HRV metrics, time averaged in the pre- and post-meal epochs are shown in Figure 5 (see also Supplemental Material Tables S2 and S3). The findings further highlight a marked divergence in baseline autonomic activity between IBS patients and healthy controls. The IBS group exhibited a substantially lower (Hedge’s g = -0.66) baseline pre-meal RMSSD (median ± MAD = 42.8 ± 16.4 ms) compared to healthy controls (71.3 ± 27.4 ms) (Figure 5a). This was accompanied by a nearly threefold elevation in the baseline Stress Index (SI) (115.3 ± 67.5 in IBS vs. 43.5 ± 24.3 in healthy controls; g = 0.72) (Figure 5d). Correspondingly, the autonomic balance ratio SI/RMSSD was 4.5x higher for the IBS cohort (6.2 ± 12.5) relative to healthy controls (0.6 ± 0.5; g = 0.59) (Figure 5g).

**Figure 5.**
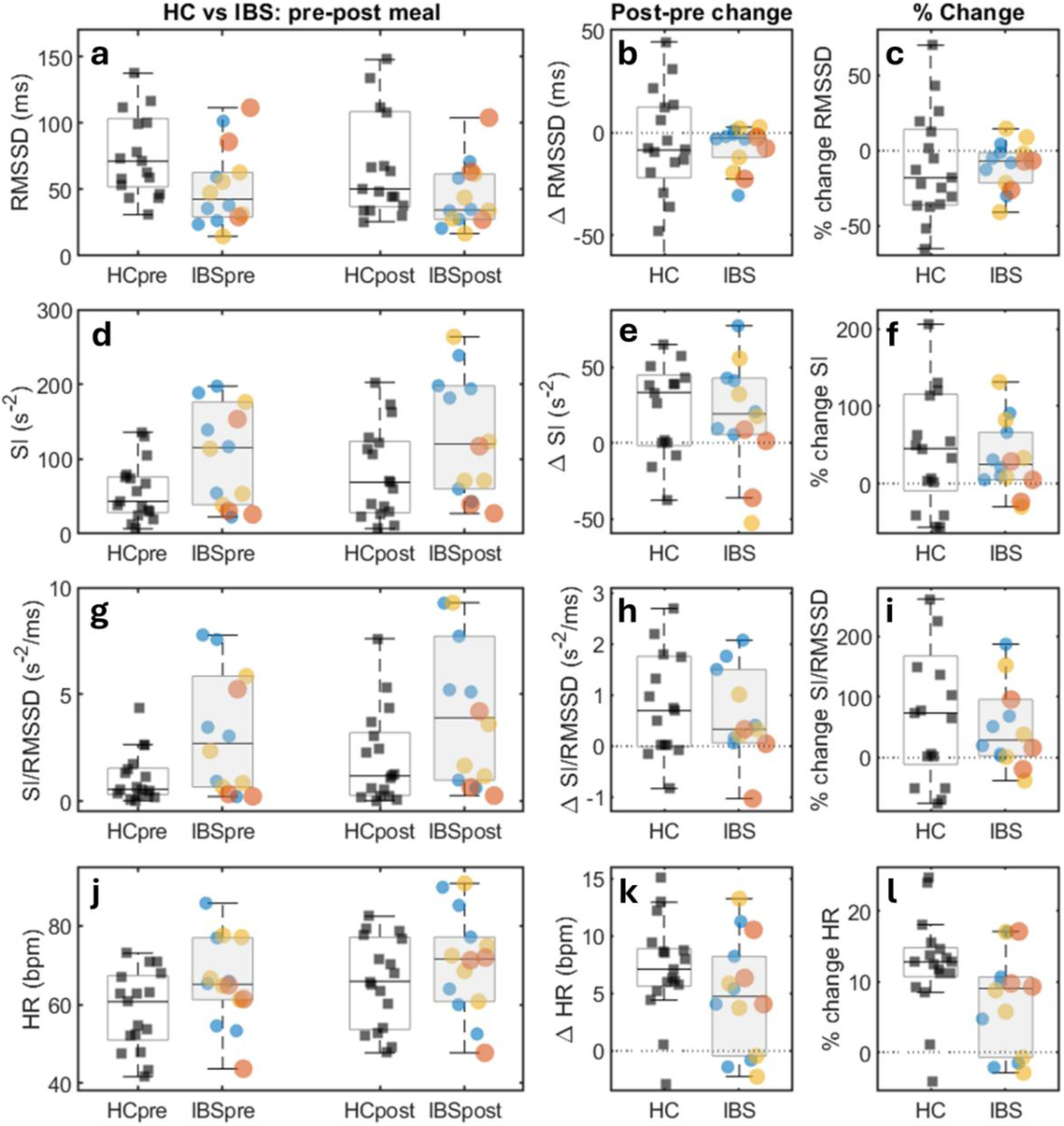
Meal response HRV metrics comparing healthy control and IBS cohorts. Rows are organized according to HRV metric. The median values in the pre and post meal epochs are plotted in the left column. Responsivity (change and percent change) are shown indicated in the middle and right columns. Each healthy control subject is represented by a black square marker. For the IBS cohort, symptom group is indicated both by the size and color of the markers (blue = none; yellow = mild; red = moderate/severe). Within each box, the central mark denotes the median and the whiskers extend to the extremum values not considered to be potential outliers.

Following the meal, a decrease in RMSSD (vagal withdrawal) and increase in SI (sympathetic activation) was observed in both IBS patients and healthy controls (Figure 5b and 5e). However, the response of both was blunted in the IBS cohort. The median meal induced changes in IBS were attenuated by 3.2x for ΔRMSSD (−2.6 ± 4.2 vs -8.4 ± 20.7 ms; Figure 5b), 1.7x for ΔSI (19.1 ± 19.9 vs 33.2 ± 31.7 s^-2^; Figure 5e); and ΔSI/RMSSD by 2.3x (0.3 ± 0.5 vs 0.7 ± 0.8 s^-2^/ms; Figure 5h). The median change in ΔHR was also blunted in IBS patients compared to controls (4.7 ± 4.3 vs 7.1 ± 1.6 bpm; Figure 5k). Taken together, group-level and subject-specific results indicate that the autonomic dysfunction in IBS is characterized by a systematic offset more so than a dysfunctional reaction to the meal challenge.

### Colonic motility - HRV dynamic trajectory

The trajectory maps illustrating dynamic coupling between colonic motility and HRV metrics are illustrated in Figure 6. More tightly bound orbits of the trajectory during the pre- and post-meal epochs, with a rapid transition between them, indicates relatively stable colonic motility-autonomic states. For RMSSD, the bounded trajectory becomes looser, with prominent deviations seen in the moderate symptom IBS group (Figure 6c and 6d). For SI, the opposite trend is observed across the IBS cohort, with the no symptoms group exhibiting the most chaotic/least bound orbit (Figure 6f).

**Figure 6.**
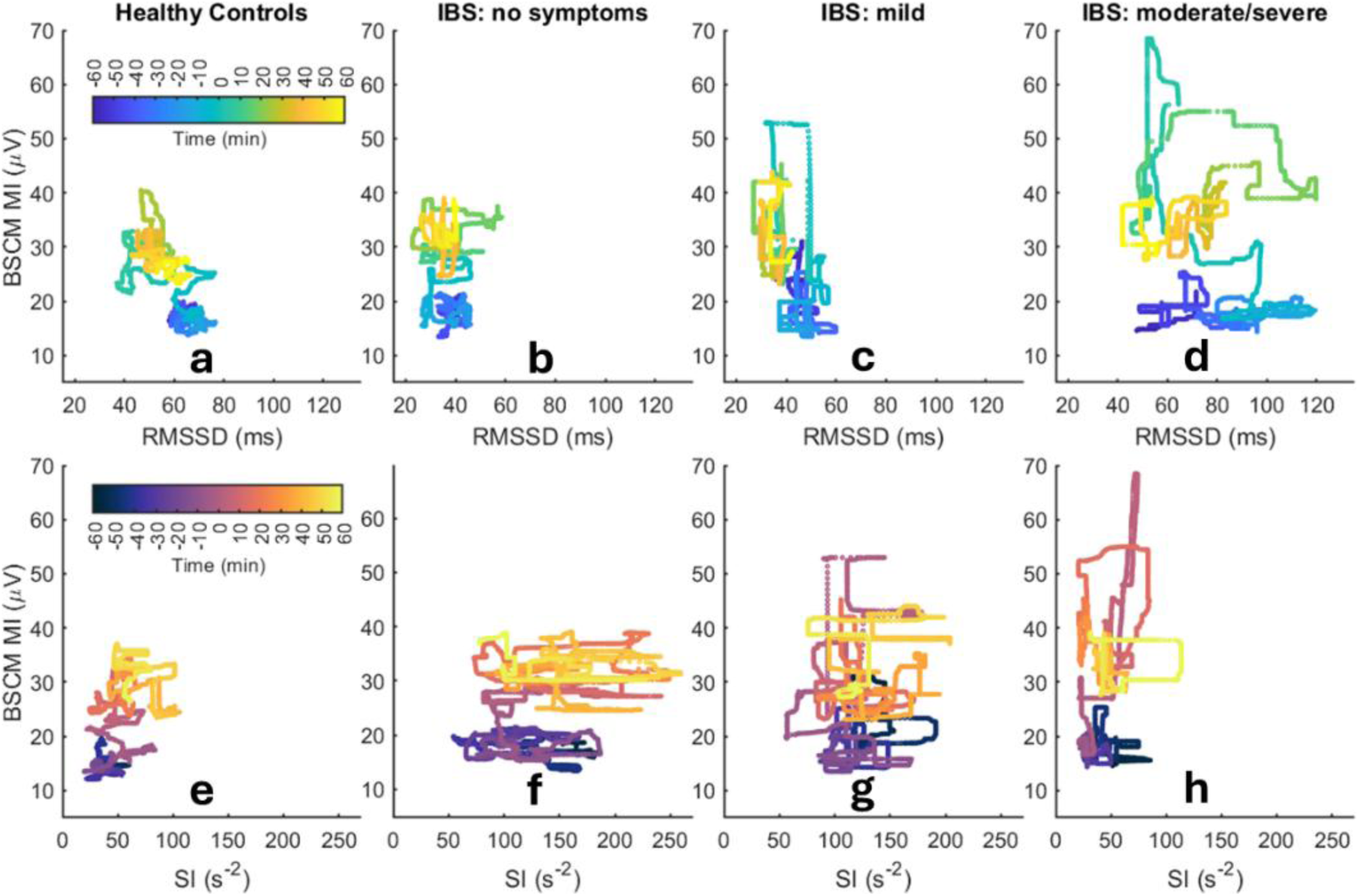
Colon motility - HRV dynamic trajectories for RMSSD (**a-d**) and SI (**e-h**). Columns are arranged by healthy controls and IBS symptom subgroup. For healthy controls, a representative trajectory from a subset (n= 6) is displayed. Trajectory color code indicates time elapsed, with meal start synced at t = 0 min.

The behavior observed in the dynamic trajectory maps is further quantified using the Euclidean distance to the centroid (Figure 7a and 7b), with smaller values indicating more compact orbits, reflecting more robust autonomic-motility regulation. Compared to healthy controls, the RMSSD-BSCM MI centroid distance *|d_c_|* for the IBS moderate symptoms was 4.0x larger than controls during the pre-meal epoch (14.8± 8.3 vs 3.7 ± 1.5) and 3.5x larger during the post meal epoch (15.2 ± 5.7 vs 4.3 ± 2.7; Figure 7a). Smaller increases of 1.2x and 1.5x were noted for the no and mild symptoms IBS group relative to controls. For SI, the SI-BSCM MI centroid distance for IBS no symptoms group was 3.3x and 3.4x larger than controls during the pre-meal (22.0 ± 12.3 vs 6.4 ± 3.0) and post-meal (31.9 ± 18.6 vs 9.7 ± 5.8) epochs (Figure 7b). The mild and moderate symptoms groups showed more modest increases of 2.0x and 1.5x, respectively.

**Figure 7.**
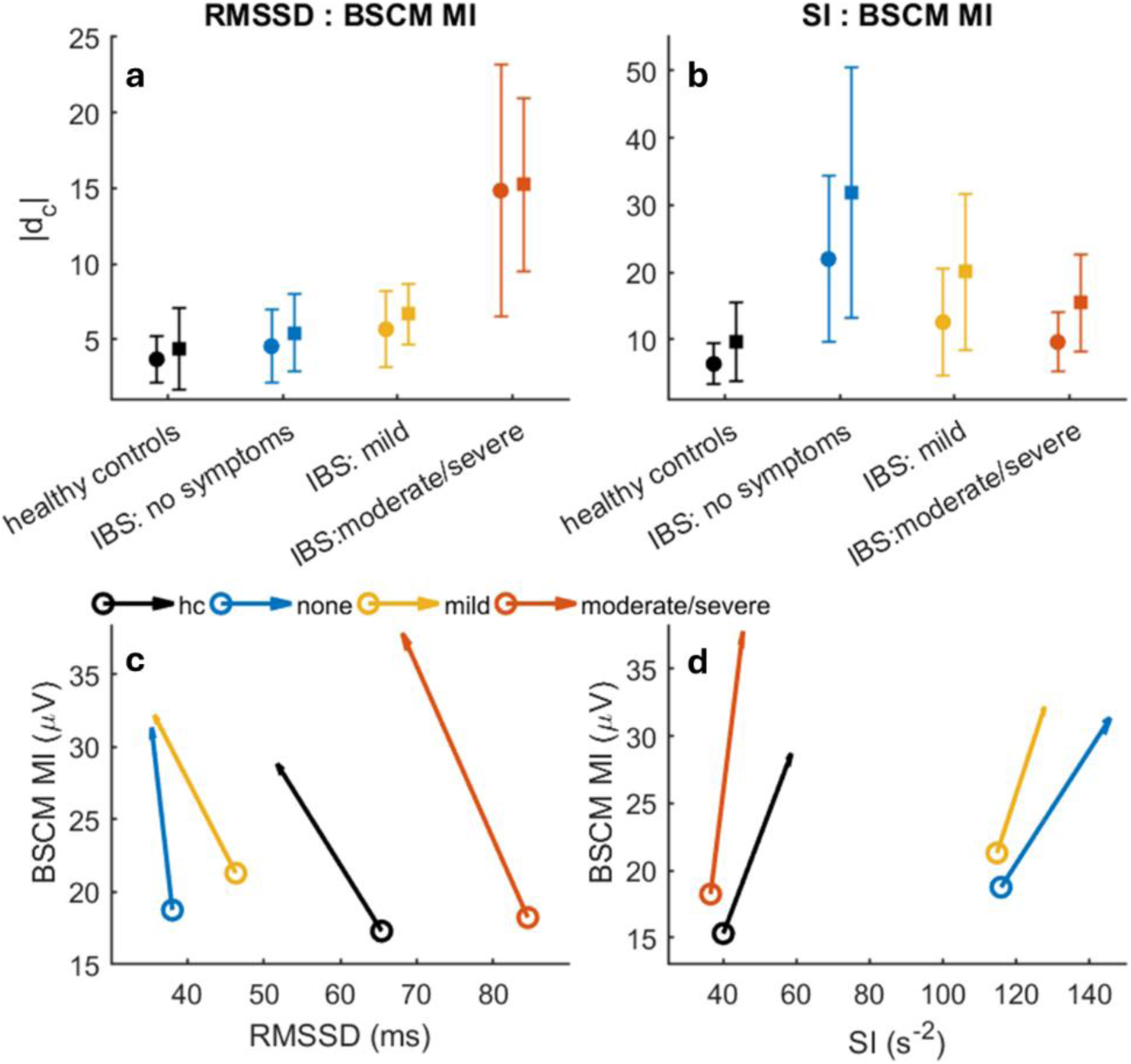
Summary metrics for HRV vs motility trajectories comparing healthy controls and IBS symptom cohorts. **a-b**: distance from centroid during premeal (circle markers) and post meal (square markers) epochs for RMSSD and SI vs motility trajectories, respectively. Data points and vertical bar indicate median + MAD. RMSSD is increasingly dysregulated (larger |d_c_|) with symptom severity, while SI shows the opposite trend. **c-d**: Vector plot highlighting meal-induced changes in HRV-motility trajectory. Vector tails (‘o’ markers) and head indicate locations for pre and -post meal epoch trajectory centroids. All groups show meal-induced vagal withdrawal (decrease in RMSSD) and sympathetic activation (increase in SI), however, the baseline condition (starting point) is vastly different, while the direction (vector angle) differs slightly across groups.

The vectors in Figure 7c-d highlight the change in trajectory stimulated by meal intake. Each vector is drawn from the centroid position of pre-meal epoch, terminating in that of the post-meal epoch. The difference in vector location highlights differences in baseline autonomic activity while their general directional alignment further indicates similarity in the dynamic response induced by the meal.

Vectors in the RMSSD-BSCM MI plane rotated anticlockwise relative to the vertical (pointing ‘northwest’) indicate vagal withdrawal (RMSSD decrease) coincides with a motility increase for healthy controls and all IBS symptom groups (Figure 7c). Between group differences were relatively modest with the IBS no symptoms group exhibiting the largest difference in trajectory axis angle (rotated anticlockwise relative to the vertical by 49.9, 12.2, 44.5, 40.0 deg, respectively). Vectors in the SI-BSCM MI plane rotated clockwise relative to vertical (pointing ‘northeast’) indicate sympathetic activation (SI increase) coinciding with motility increase for all groups (Figure 7d). The IBS moderate symptoms group exhibited the largest difference in trajectory angle (24.2 deg vs 54.2, 67.1, 50.1 deg for healthy controls, IBS no and mild symptoms, respectively).

### Symptoms, motility, and autonomic function: case studies

Two case studies illustrate how BSCM and HRV may be used to elucidate their role in symptom genesis. The autonomic balance ratio SI/RMMSD is utilized as a single metric that captures changes in either of these metrics (but does not resolve which one).

The first patient (Figure 8) experienced multiple bouts of gas, pain, cramping, culminating in strong urge to defecate toward the end of the study. Each symptom episode correlated with a peak in colonic motility, only one of which was correlated to a large spike in autonomic activity (Figure 8b). The initial episode of gas and cramping (t ∼ 20 min) correlated to a strong increase in MI but no apparent change in SI/RMSSD. The second bout of pain and cramping (t ∼ 55 min) correlated to a strong MI increase, with a prominent autonomic activity peak indicating a shift toward sympathetic dominance occurring immediately after MI peak. Lastly, strong urge (t∼78min) was again correlated to a peak in MI, with a small amplitude transient decrease in SI/RMSSD. While causality cannot be discerned at present, these results suggest colonic motility alone can serve as the primary mechanism of symptom genesis.

**Figure 8.**
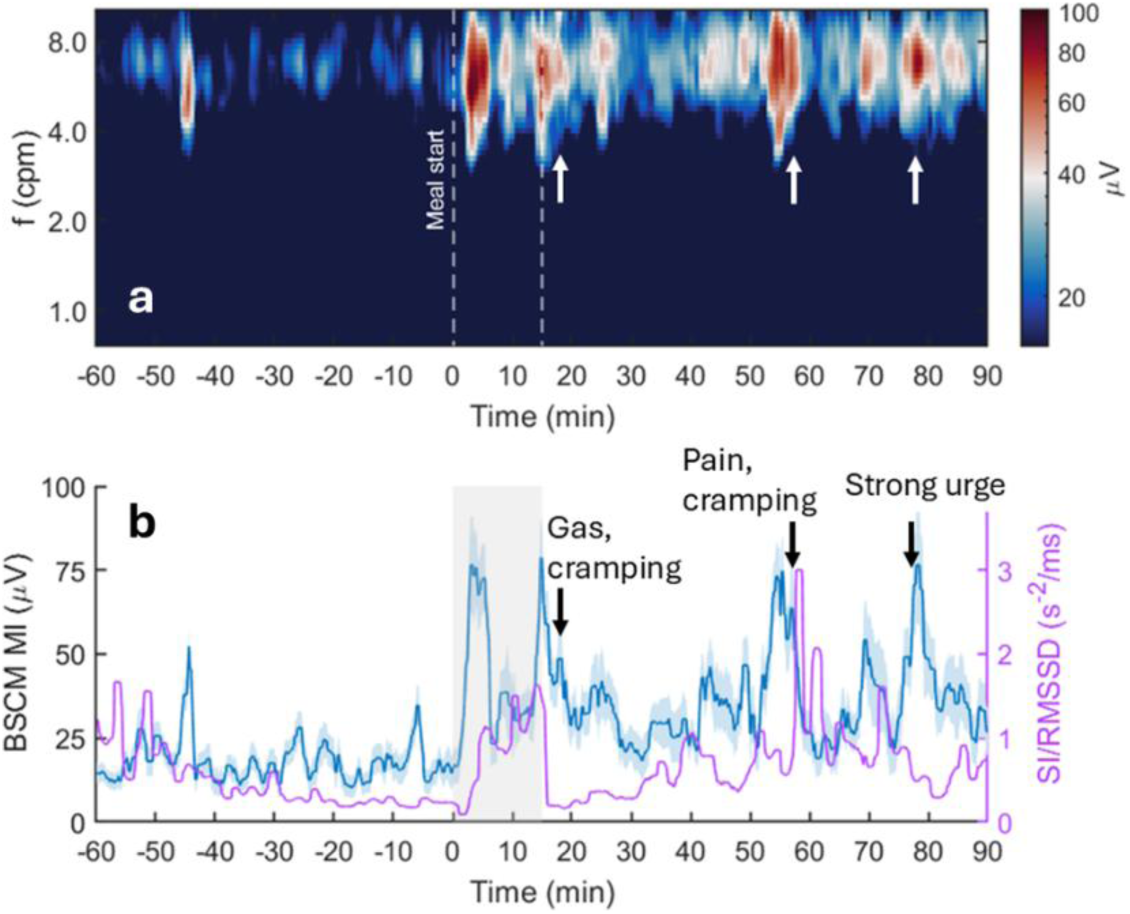
Relationships between colon motility, autonomic function, and patient symptoms. **a**: BSCM spectrogram for the 4-10 cpm passband. (Note: log color scale). White arrows mark the occurrence of patient-reported symptoms. **b**: BSCM MI and SI/RMSSD indicating motility and autonomic function in relation to patient-reported symptoms.

The second case study (Figure 9) experienced gas and bloating immediately after eating, following by two bouts of defecation urge, the latter of which was reported to intense and painful. The gas and bloating phase coincides with meal-induced increase in motility in the 4-10 cpm band and a subsequent increase in SI/RMSSD indicating a sympathetic shift in ANS activity (Figure 9a-b). Its possible that physical distress due to symptoms is driving the sympathetic shift. The first report of urge (t ∼ 45 min) coincides with a pulsatile decrease in SI/RMSSD but no pronounced change in motility index. In contrast, the second strong bout of urge (t∼70 min) shows no notable autonomic shift but does coincide with a marked transient motility increase (local peak in BSCM MI).

**Figure 9.**
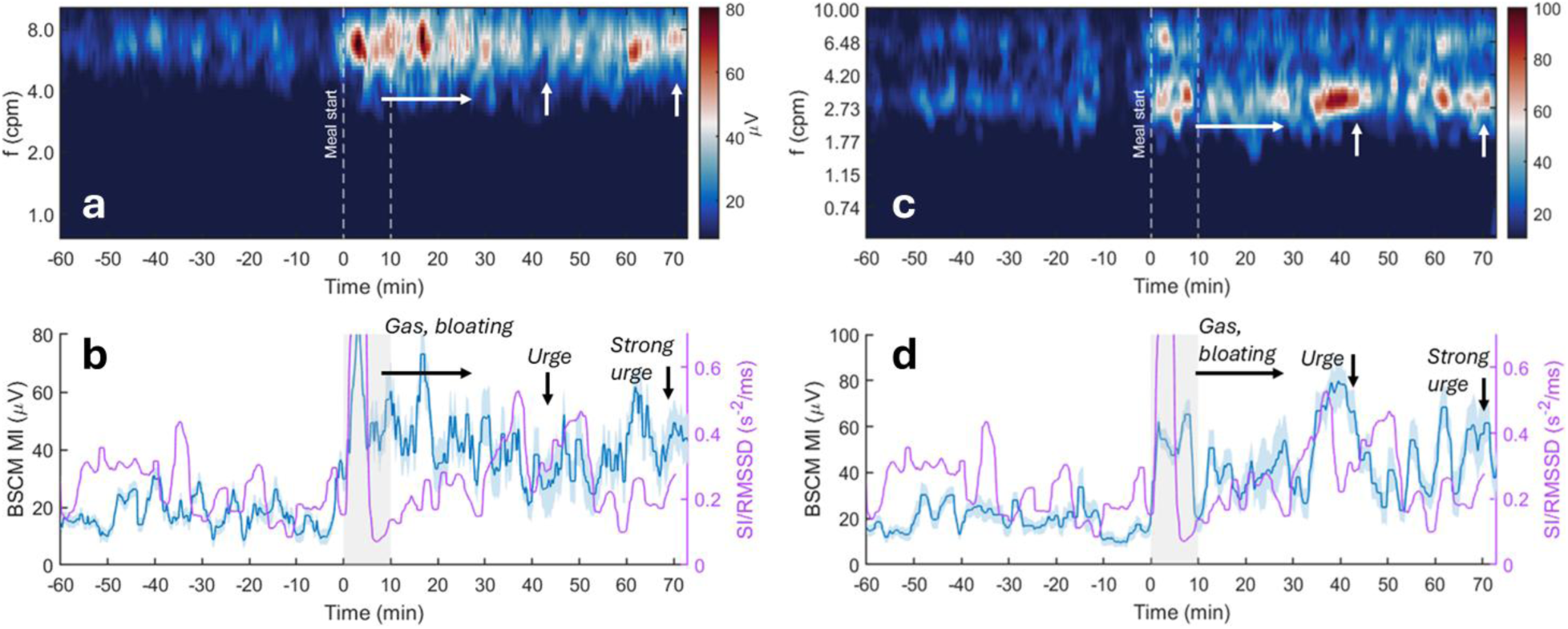
Case study of a patient experiencing moderate/severe symptoms with relation to colonic motility and autonomic function. **a-b**: 4-10 cpm bandpass filtered motility data; **c-d**: 0.5-10 cpm passband motility data. Format for each panel is the same as Figure 8.

Additionally, analyzing colonic motility with signals filtered in the 0.5-10 cpm passband (Figure 9c-d) reveals strong rhythmic ∼3.5 cpm motilty coincident with the first bout of urge. This could be a manifestation of high-amplitude propagating contractions (24, 35, 39). The second bout of urge also coincindes with a prominent MI increase in this wider band, which may be a manifestation of multiple simultaneously active motor patterns driven by distinct ICC networks (40, 41).

## DISCUSSION

Predominant theories for the pathogenesis of IBS include visceral hypersensitivity and abnormal colonic motility, with a focus on an integrated model of disordered gut-brain interaction (42, 43).

Whereas the literature investigating autonomic dysfunction in IBS patients is extensive (11, 14), only a few published reports analyze concurrent measurements of colonic motility in health (24, 39) and disease (22, 44). The present work establishes technical feasibility and practical utility of a novel non-invasive method for investigating the interplay between colonic motility, ANS function, and IBS symptom genesis.

This small cohort pilot study revealed several preliminary but intriguing hypothesis-generating findings linking colonic motility and autonomic function. Meal-induced colonic motility was strongly increased in patients experiencing moderate to severe abdominal symptoms, indicating hypermotility may play a role in driving IBS symptoms. In the same symptom group the vagal-motility trajectory was prominently dysregulated, in contrast to the sympathetic-motility trajectory which remained tightly regulated. This suggests disruption of the vagal input modulating normal motility. Finally, the highest baseline sympathetic activity and most dysregulated SI-motility trajectory was observed in IBS patients reporting no symptoms, and the converse was true for the most symptomatic patients. Increased perception of intestinal wall stretching correlated to sympathetic activation was previously proposed as primary symptom-generating mechanism, the present results are contrary to this hypersensitivity model. Sympathetic input nominally inhibits some types of colonic motility (24, 39), hence one working hypothesis is that sympathetic drive acted to suppress motility, hence symptoms.

Consistent with the majority of previous studies, our IBS patients exhibited decreased parasympathetic and increased sympathetic activity relative to healthy controls (11, 14, 43). Our patients also exhibited an attenuated ANS response to a meal, consistent with blunted vagal and sympathetic reactivity to visceral stressors like sigmoidoscopy (45) and colorectal distention (46). These findings align with muted SI/RMSSD meal response observed here. Furthermore, the highly dysregulated vagal activity in our high-symptom group is consistent with the profoundly altered HRV profiles identified in severe IBS-D subgroups (15).

A previous manometry study linked postprandial colonic motility to vagal withdrawal consistent with our non-invasive findings, although the same study found no correlation between symptom severity and HRV parameters (22). Conversely, studies in healthy volunteers associated meal-induced HAPCs with transiently increased parasympathetic (RMSSD) and decreased sympathetic (SI) activity (24, 39) (Yuan and Huizinga 2020; Ali & Chen 2021). The apparent discrepancy from our motor-autonomic association likely stems from our BSCM signal processing pipeline optimized to identify lower amplitude cyclic motor patterns, rather than HAPCs (34).

High-resolution manometry has previously characterized a diminished cyclic motor pattern and increased antegrade propagating contractions in high-symptom IBS-D (20). As HAPCs have been temporarily associated with episodes of pain in other earlier studies (47), we hypothesize that these events may underly the marked BSCM motility increase observed in our high symptom IBS group. This hypothesis may be tested in the future, as BSCM has recently been shown to identify bisacodyl-induced HAPC by modifying the signal processing parameters (35).

This study has several limitations. The small sample size necessitates a larger cohort to definitively characterize differences in colonic motility and autonomic function between IBS and healthy controls. HRV is an indirect measure for autonomic function. There are no accepted reference ranges for metrics like RMSSD or SI in IBS patients (11), and baseline sympathetic dominance may reflect anticipatory meal anxiety rather than chronic dysautonomia. Medication use was not restricted, and meals were not standardized. While allowing patients to choose familiar foods may have reduced meal stress, these factors could still influence the results. This study stratified data by subjective severity during the meal test. Utilizing a standardized system for recording real-time symptom scores could help to improve grouping (48, 49).

The cohorts in this study were not sex and aged matched. The gender (assigned at birth) of IBS cohort was consistent with ∼3:2 (F:M) ratio reported for the USA and the with a 2:1 ratio for those seeking medical care (50, 51). HRV parameters (RMSSD or HF) are typically higher in women and decrease with age (22, 45), while sympathetic markers (SI or LF) increase with age (22). Future studies should also account for the phase of menstrual cycle at the time of the experiment, as IBS symptoms vary with hormonal phases. (52, 53).

Finally, while this study focused on the colon, gastric and small bowel dysmotility can also contribute to IBS symptoms (54). Expanding the electrode array to simultaneously measure contributions from all three organs may help to isolate organ specific association with IBS symptoms (55).

In summary, we have presented a new technique based on cutaneous electrical recordings as a painless, easy-apply, low-resource requirement, cost-effective, tool for investigating colonic motility and autonomic activity. This scalable approach provides a framework for identifying the mechanisms of symptom genesis in IBS and other colonic disorders, such as refractory chronic constipation. This work also introduced dynamic motility-HRV trajectory plots for visualizing and quantifying motor events in relation to autonomic function. This approach offers a framework for characterizing disorders of gut-brain interaction, providing a mechanistic basis for optimizing interventional treatment strategies. This novel methodology could be applied to larger cohort studies to help characterize and elucidate the associations and causal relationships between autonomics and motility measures in IBS, as well as a range of other gastrointestinal disorders, such as functional dyspepsia and refractory chronic constipation. While the novel methodology presented here focused on measurement of colonic motility, it could also be easily adapted and extended to multi-modal studies of gastric motility in relation to autonomic function (56).

## DATA AVAILABILITY

Data will be made available upon reasonable request to the corresponding author.

## SUPPLEMENTAL MATERIAL

Supplemental Tables S1-S4 Supplemental Figs. 1-2

## Data Availability

All data produced in the present study are available upon reasonable request to the authors

## ACKNOWLEDGMENTS

We thank Sibylle Van Hove for fruitful discussions regarding analysis of altered motility and HRV parameters in the context of symptom severity.

## AUTHOR CONTRIBUTIONS

Conceived and designed research (JCE, LP, JG), performed experiments (LP, NG, JG) analyzed data (JCE, LP, NG, JG), interpreted results of experiments (JCE, LP, PGD), prepared figures (JCE), drafted manuscript (JCE, LP, PGD), edited and revised manuscript (JCE, LP, PGD), approved final version of manuscript (JCE, LP, JG, NG, PGD).

## SUPPLEMENTAL MATERIAL

### Motility Index metrics: Summary Statistics for meal-response parameters

**Table S1.** Group averaged BSCM motility metrics. MRR = meal response ratio. APD = amplitude percent difference. Data are presented as median statistics and normal distribution parameters.

| Metric | Group | Median ± MAD | Mean ± S.D. |
| --- | --- | --- | --- |
| **MRR** | Healthy control | 1.86 ± 0.49 | 1.82 ± 0.61 |
| | IBS: no symptoms | 1.58 ± 0.21 | 1.61 ± 0.34 |
| | IBS: mild | 2.35 ± 0.71 | 2.04 ± 0.86 |
| | IBS: moderate/severe | 2.92 ± 0.50 | 2.67 ± 0.66 |
| **APD (%)** | Healthy control | 52.7 ± 25.3 | 56.5 ± 30.6 |
| | IBS: no symptoms | 51.2 ± 14.73 | 58.0 ± 18.7 |
| | IBS: mild | 36.4 ± 24.07 | 49.0 ± 28.5 |
| | IBS: moderate/severe | 102.9 ± 21.0 | 102.1 ± 31.0 |

### HRV metrics Summary Statistics

**Table S2.** HRV parameters for meal epochs and the meal induced change. Each cell is formatted as median statistics (Median ± MAD); normal distribution parameters (Mean ± SD). Statistically significant differences (based on 95% CI) between healthy controls and IBS are denoted by *. Units: RMMSD (ms), SI (s^-2^), SI/RMSSD (s^-2^/ms), HR (bpm).

| Metric & Cohort | Pre-meal | Post-meal | Post-Pre Change | Change (%) |
| --- | --- | --- | --- | --- |
| **RMSSD (Healthy)** | 71.3 ± 27.4;<br>89.8 ± 70.6 | 50.4 ± 16.2;<br>82.0 ± 77.2* | -8.4 ± 20.7; -<br>7.8 ± 28.8 | -18.1 ± 19.6; -<br>9.8 ± 35.2 |
| **RMSSD (IBS)** | 42.8 ± 16.4;<br>51.7 ± 29.9 | 34.6 ± 11.6;<br>44.8 ± 24.2* | -2.6 ± 4.2;<br>-7.0 ± 10.3 | -7.0 ± 8.5;<br>-9.6 ± 15.5 |
| **SI (Healthy)** | 43.5 ± 24.3;<br>57.0 ± 39.1 | 69.1 ± 43.9;<br>81.3 ± 61.0 | 33.2 ± 31.7;<br>24.3 ± 50.5 | 45.2 ± 68.8;<br>58.6 ± 107.4 |
| **SI (IBS)** | 115.3 ± 67.5;<br>141.6 ± 164.9 | 120.3 ± 75.4;<br>168.2 ± 178.5 | 19.1 ± 19.9;<br>26.6 ± 48.9 | 24.7 ± 19.8;<br>32.8 ± 45.2 |
| **SI/RMSSD (Healthy)** | 0.6 ± 0.5;<br>1.1 ± 1.2 | 1.2 ± 1.1;<br>2.0 ± 2.2 | 0.7 ± 0.8; 0.9<br>± 2.1 | 73.0 ± 74.8;<br>153.8 ± 319.7 |
| **SI/RMSSD (IBS)** | 2.7 ± 2.4;<br>6.2 ± 12.5 | 3.9 ± 3.1;<br>7.0 ± 12.4 | 0.3 ± 0.5; 0.8<br>± 2.1 | 28.3 ± 33.7;<br>62.0 ± 92.3 |
| **HR (Healthy)** | 60.7 ± 8.4; 58.4<br>± 10.0 | 65.8 ± 11.9;<br>65.5 ± 11.4 | 7.1 ± 1.6; 7.2<br>± 4.3 | 12.8 ± 1.8; 12.4<br>± 6.9 |
| **HR (IBS)** | 65.1 ± 7.2; 65.6<br>± 11.1 | 71.6 ± 9.2; 70.5<br>± 12.9 | 4.7 ± 4.3; 4.8<br>± 4.9 | 9.0 ± 6.2;<br>7.3 ± 7.1 |

**Table S3.**
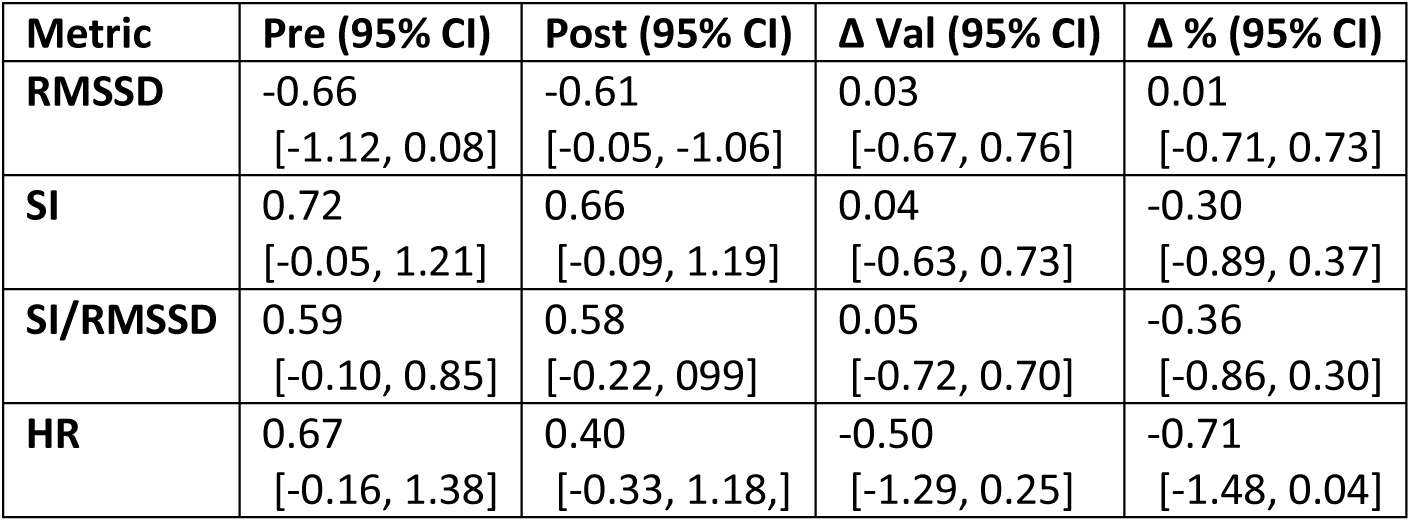
Cohen’s D effect sizes and confidence intervals for HRV parameters of IBS relative to controls. A negative value indicates a decrease for IBS relative to controls; a positive value indicates a positive shift for IBS relative to controls. The first two columns list pre-meal epoch (‘Pre’) and post-meal epoch (‘Post’) values. The last two columns indicate responsiveness to the meal as value percent difference post- vs pre-meal (‘Δ Val’); and percent difference (‘Δ %’). The results indicate moderate to significant baseline shifts in HRV between IBS groups and controls. However, owing to the large variance in the healthy controls, the small Hedge’s-g values would suggest responsivity is not significantly different.

**Figure S1.**
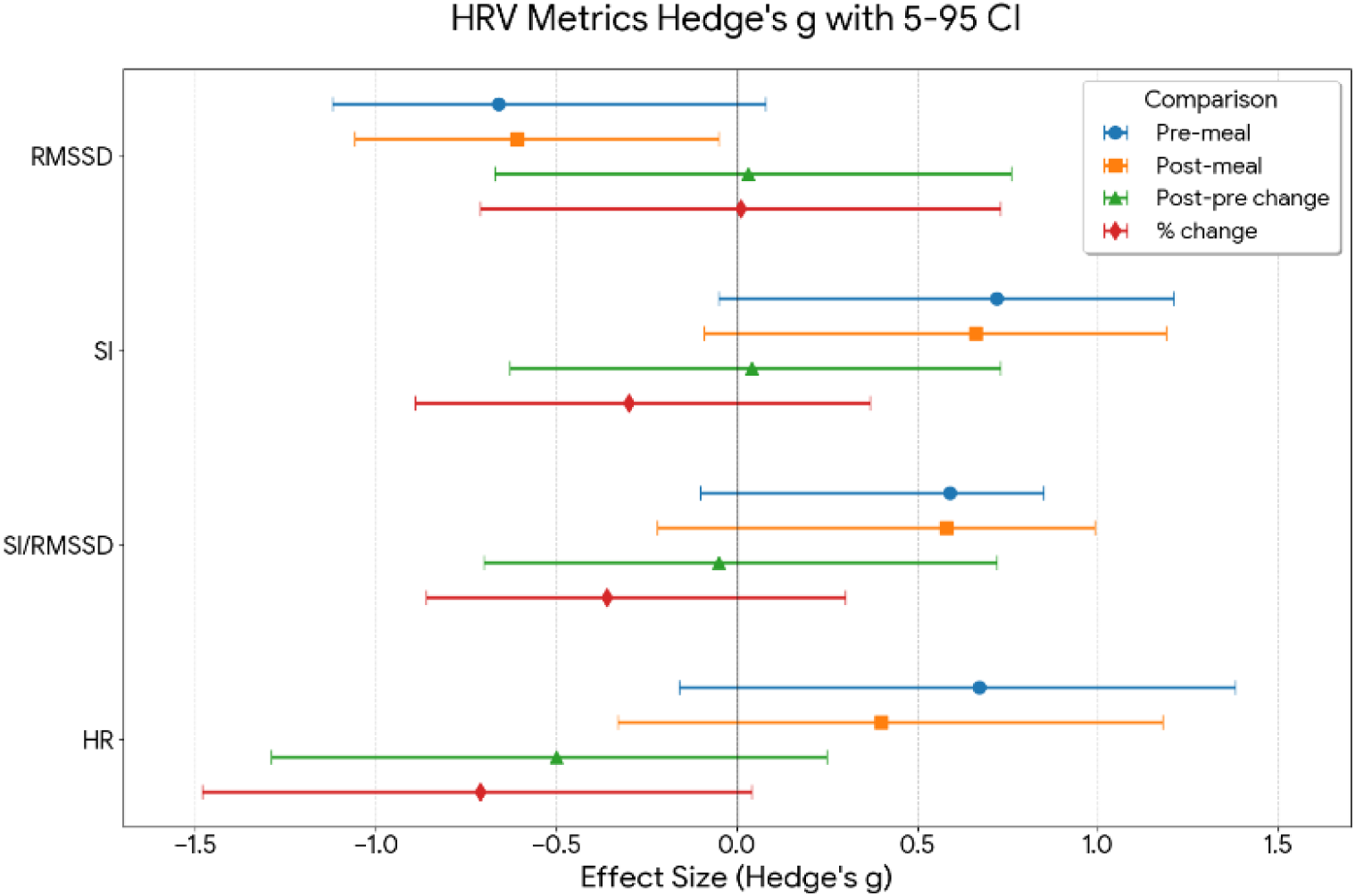
Forest plot summary visualizing data in Table S3.

### Dynamic trajectory plots: healthy controls subset analysis

Given the larger cohort size of healthy controls compared to IBS subgroups, we wondered whether the more constrained motility-HRV trajectories resulted from an averaging effect, as opposed to a genuine physiological difference. To address this, we computed trajectories for a comparable -sized subset of controls (*n_sub_* = 6). We generated *n_r_* = 30 realizations using randomly selected permutations and averaged those 6 results together to generate a single motility-HRV trajectory.

Next, we computed the bivariable kernel smoothed density maps of the trajectories. Doing so facilitated comparison of trajectories within and between groups by computing the Pearson correlation coefficients of the trajectories (treated as 1-dimensional vectors).

The self-similarity of the 30 control subset trajectories was quantified by computing the correlation for all possible (30 x 29) pairs. Each of the 30 controls subsets were also compared against the full cohort of controls (n = 22) and to each of the IBS symptoms cohorts (i.e. 30 x 1 correlation coefficients were computed for each group comparison).

The results of this analysis are shown in Figure S3 and Table S4. Results are displayed for both RMSSD-MI (blue) and SI-MI (red). For RMSSD, the correlation within controls subsets and between subsets and the full control cohort were similar. The controls subsets versus IBS no and mild symptoms trajectories were substantially different, with little overlap in their distributions (manifested as shift downward in correlation coefficient). While the moderate/severe symptoms were very similar to controls, this is an artifact deriving from the wide area coverage of the unconstrained trajectory for the IB severe group (See Table S5). For SI, controls subsets were internally consistent but were substantially different compared to all IBS symptoms groups. In summary, this analysis shows a negligible averaging effect from having a larger controls cohort and offers support of the conclusion that there are substantial physiological differences between healthy controls and IBS, manifested as differences in their autonomics-motility trajectories.

**Figure S2.**
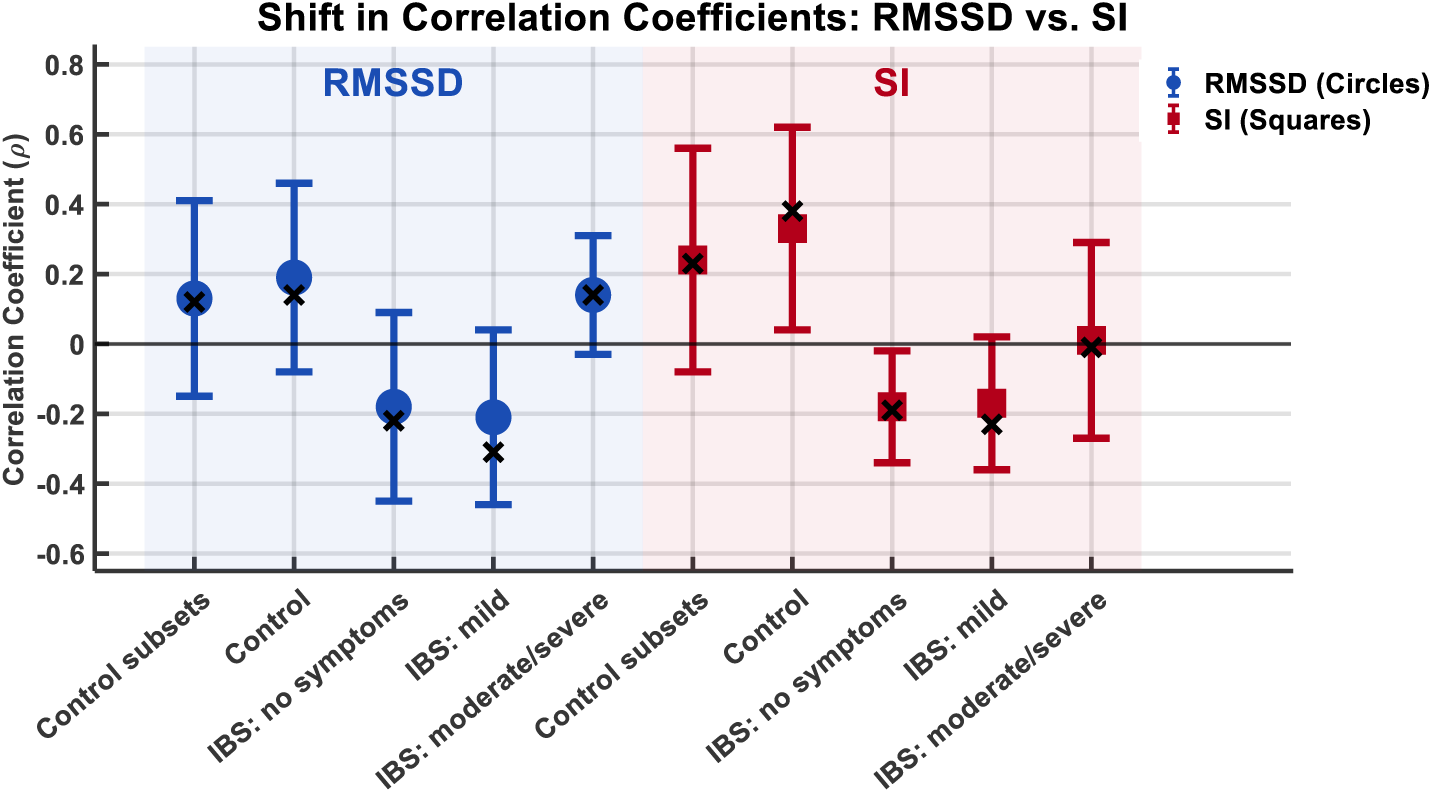
Correlation of HRV-motility trajectories comparing control subsets to the 4 main groups analyzed throughout this work (full cohort of controls + 3 IBS groups). The data points (circles and squares) indicate the mean of the correlation coefficient distribution, while ‘x’ marks the median. The extent of the vertical bars indicates the 5-95% confidence interval.

**Table S4:** Correlation coefficient distribution parameters resulting from controls subcohort trajectory analysis.

|  | RMSSD |  |  | SI |  |  |
| --- | --- | --- | --- | --- | --- | --- |
|  | Median | Mean | Std Dev | Median | Mean | Std Dev |
| Control subsets | 0.12 | 0.13 | 0.28 | 0.23 | 0.24 | 0.32 |
| **Control full cohort** | 0.14 | 0.19 | 0.27 | 0.38 | 0.33 | 0.29 |
| **IBS no symptoms** | -0.22 | -0.18 | 0.27 | -0.19 | -0.18 | 0.16 |
| **IBS mild symptoms** | -0.31 | -0.21 | 0.25 | -0.23 | -0.17 | 0.19 |
| **IBS moderate/severe** | 0.14 | 0.14 | 0.17 | -0.01 | 0.01 | 0.28 |

**Table S5:** Data Counts used for density map correlation. Summary of observation of valid overlapping elements (with non-zero value) in the kernel smoothed density maps used for correlation computation across the study groups. The image grid was 200 x 200 hence 40,000 is the maximum possible. Only a small fraction of overlap was present in each case, however high symptom group showed the highest overlap in RMSSD, reflecting the unconstrained trajectories.

|  | **RMSSD** |  |  | **SI** |  |  |
| --- | --- | --- | --- | --- | --- | --- |
|  | **Median** | **Mean** | **Std Dev** | **Median** | **Mean** | **Std Dev** |
| **Control subsets** | 1463 | 1433 | 483 | 1257 | 1225 | 352 |
| **Control full cohort** | 1235 | 1172 | 274 | 850 | 828 | 171 |
| **IBS no symptoms** | 616 | 682 | 430 | 1065 | 1177 | 564 |
| **IBS mild symptoms** | 1092 | 1212 | 573 | 945 | 1106 | 526 |
| **IBS moderate/severe** | 1974 | 2005 | 657 | 938 | 954 | 353 |

### Colonic motility in the 0.5 – 10 cpm pass band

We also analyzed colonic motility in a wider passband of 0.5 – 10 cpm. The motivation for doing using this alternate frequency band was: 1) It was used to detect meal-induced colonic cyclic motor patterns in a previous report (31); 2) It has recently been shown to be optimal for detecting another characteristic type of motility, high-amplitude propagating contractions, which are known to precede defecation (35); and 3) colonic motility at 2-8 cpm is known to occur (57, 58). While gastric activity operating in a narrow band around 3 cpm also falls within this range, colonic activity is believed to dominate, as the electrode array is relatively distant from the stomach and centered directly above the descending and rectosigmoid colon. The results for the 0.5-10 cpm band are similar to that of the 4-10 cpm. They indicate a dramatic increase in motility for the IBS moderate/severe symptoms group, while the no and mild symptoms groups exhibit modest to negligible differences compared to controls.

**Figure S3.**
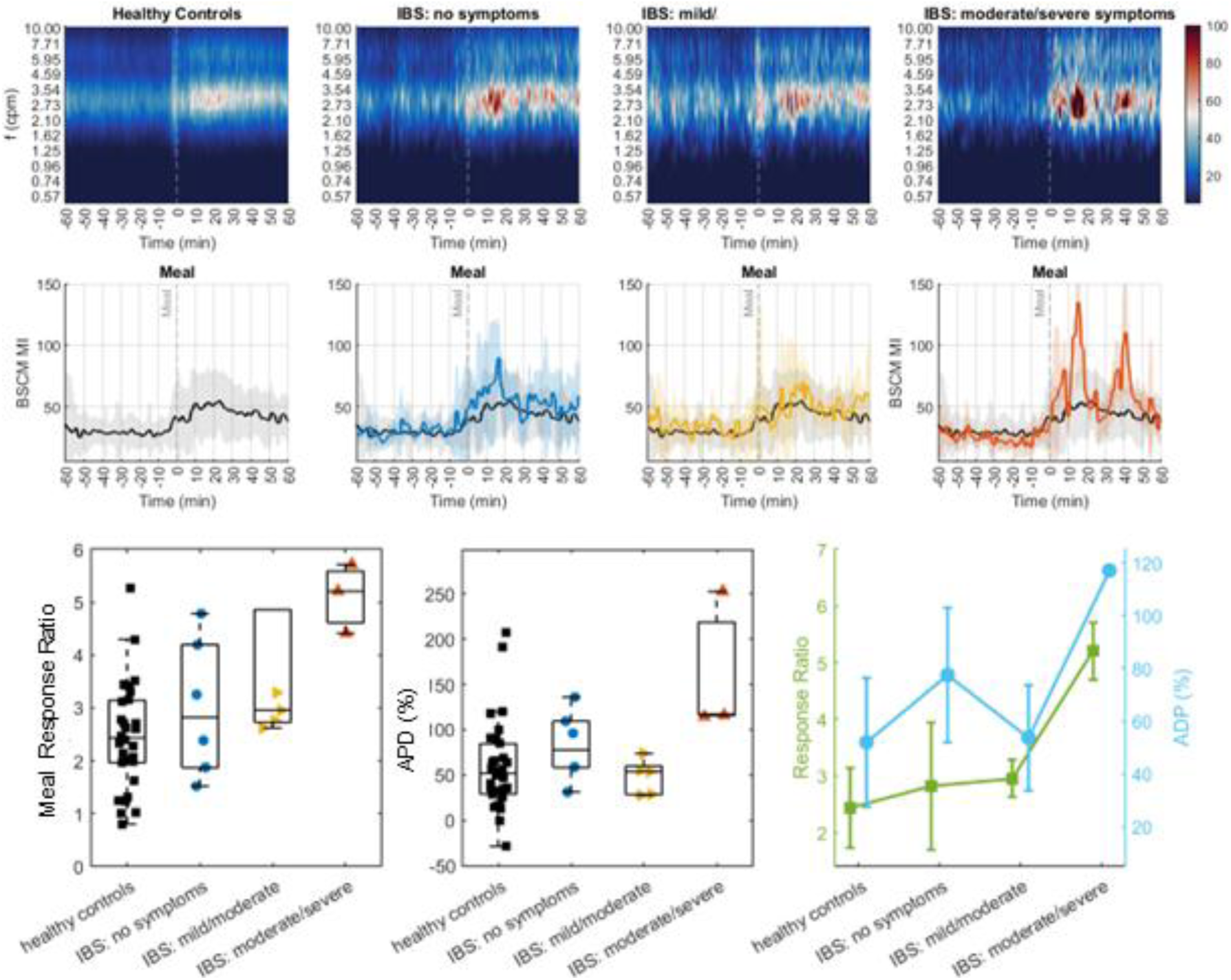
Colonic motility metrics derived from the 0.5 – 10 cpm band comparing IBS symptom groups to healthy controls. Top row: BSCM spectrograms for healthy controls and IBS symptom groups in order of increasing symptom severity. Middle row: Motility Index for each of the 4 groups. The black trace (controls) is presented across all panels to facilitate comparisons to IBS symptom groups. The moderate/severe symptoms group exhibits large amplitude deviations compared to controls. Bottom row: Meal response ratio and Amplitude Percent Difference (ADP %).

